# A parametric bootstrap approach for computing confidence intervals for genetic correlations with application to genetically-determined protein-protein networks

**DOI:** 10.1101/2023.10.24.23297474

**Authors:** Yi-Ting Tsai, Yana Hrytsenko, Michael Elgart, Usman Tahir, Zsu-Zsu Chen, James G Wilson, Robert Gerszten, Tamar Sofer

## Abstract

Genetic correlation refers to the correlation between genetic determinants of a pair of traits. When using individual-level data, it is typically estimated based on a bivariate model specification where the correlation between the two variables is identifiable and can be estimated from a covariance model that incorporates the genetic relationship between individuals, e.g., using a pre-specified kinship matrix. Inference relying on asymptotic normality of the genetic correlation parameter estimates may be inaccurate when the sample size is low, when the genetic correlation is close to the boundary of the parameter space, and when the heritability of at least one of the traits is low. We address this problem by developing a parametric bootstrap procedure to construct confidence intervals for genetic correlation estimates. The procedure simulates paired traits under a range of heritability and genetic correlation parameters, and it uses the population structure encapsulated by the kinship matrix. Heritabilities and genetic correlations are estimated using the close-form, method of moment, Haseman-Elston regression estimators. The proposed parametric bootstrap procedure is especially useful when genetic correlations are computed on pairs of thousands of traits measured on the same exact set of individuals. We demonstrate the parametric bootstrap approach on a proteomics dataset from the Jackson Heart Study.

## Introduction

Genetic correlation measures the relationship between a pair of traits through their shared genetic variability (1). It is a related concept to heritability, which measures the overall genetic contribution to a trait (2). Specifically, genetic correlation is defined as the correlation between the genetic effects of two traits. It can be estimated using individual-level data, or using summary statistics from genome-wide association studies (GWAS) (3). Scientific papers studying genetic architecture of health and behavioral phenotypes now routinely report genetic correlation estimates between phenotypes, sometimes as a step preceding follow up analysis, e.g. with polygenic risk scores or Mendelian randomization analyses (4–7). Genetic correlations are further being studied at the local genomic region level (local genetic correlations), or stratified by genetic annotations, to localize sources of shared genetic underpinning of phenotypes (8–12).

Methods for estimating heritability and genetic correlations based on summary statistics from GWAS (3,13,14) became popular in recent years due to their computational tractability and the access to many phenotypes that were interrogated in GWAS by the research community. However, in diverse populations and in small datasets it is still preferable to estimate heritabilities and genetic correlations using individual-level data, rather than based on GWAS summary statistics (15). Methods using individual-level data typically rely on an underlying linear mixed model (LMM) formulation, where a genetic relationship matrix is used to model the relationship, or degree of similarity, between the phenotype levels of different individuals (16,17). When estimating genetic correlation between two phenotypes, a bivariate normal model is usually used. Common algorithms for estimating heritability and genetic correlation include Restricted Maximum Likelihood (REML)-based normal likelihood models (18), and method of moment estimators such as the Haseman-Elston approach (15). Estimation of standard errors (SEs), confidence intervals (CIs), and p-values, often relies on asymptotic normal approximation. However, both heritability and genetic correlations have a limited support: heritability is bounded on the [0,1] interval, and genetic correlation on the [-1,1] interval. This means that asymptotic normal approximation may not be appropriate when estimates are close to the boundary of the parameter space, and the problem is more severe with smaller datasets. Previous publications addressed the problem of confidence interval estimation in the context of heritability (19,20), but, although the distribution of genetic correlations has been studied (21–23), methods for confidence interval computation in the era of large-scale genomic studies have not been as developed. Notably, we previously proposed a Fisher’s transformation-based approach and a blocked bootstrap, relying on resampling from the data, by blocks of related individuals (15). The blocked bootstrap worked better than the Fisher’s transformation approach, but was computationally more intensive and we therefore only allowed for a small number of resamples, limiting the potential coverage of the confidence intervals as well as application at scale (i.e., for millions of traits). Here, we build on a prior work by Schweiger et al. (19), in the context of heritability. We expand their parametric bootstrap test-inversion method which eliminates the dependency on asymptotic approximation.

In this paper, we develop a parametric bootstrap approach to construct CIs for genetic correlations to better model the unknown distribution of genetic correlations. The procedure requires simulating pairs of phenotypes using existing correlation structure between individuals in a given dataset, based on sets of values of heritabilities and genetic correlation between the phenotypes. The results from the simulation study are used to construct CIs for the genetic correlation parameter based on triplets of estimated heritabilities and genetic correlation of a pair of phenotypes, using the conditional empirical probability mass function (PMF) of the genetic correlation parameter. We demonstrate and compare, through simulations, the performance of two variations of the parametric bootstrap procedure, and further compare them with construction of CIs based on the Fisher’s transformation of the estimated genetic correlation, and estimated standard errors (SEs) of the correlation parameter from asymptotic normal assumption on restricted maximum likelihood estimates. Despite being a resampling method, typically requiring many computations and thus computationally costly, our approach is very useful when estimating genetic correlations between thousands of traits measured on the same dataset, because the simulation study used to construct PMFs is performed once and may be used many times. Thus, we demonstrate the application of the parametric bootstrap approach to study genetic correlations between a high-dimensional set of proteins and to develop protein-protein networks based on the genetic correlations estimated in the Jackson Heart Study dataset.

## Methods

### Linear Mixed Model (LMM) formulation

Let ***y*** be an *n* × 1 phenotype outcome vector and ***X*** be an *n* × *p* matrix containing values of *p* covariates measured on *n* participants. Let ***e*** be an *n* × 1 vector of residuals, or errors, which we assume are potentially correlated across participants due to shared genetic effects. Suppose that the *n* × *n* matrix ***K*** models the genetic relationship between individuals, such that its *i,j* entry *k*_*i,j*_ is, for example, (twice) the kinship coefficient between individual *i* and *j*, and is equal to *k*_*j,i*_ = *k*_*i,j*_ (i.e., this is a symmetric matrix). Note that genetic relationship could be estimated by various quantities (24), without loss of generality, we here assume that we use a kinship matrix using identity by descent estimates. Following standard linear mixed model formulation of heritability, we model the outcome as

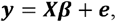

where ***β*** are the regression coefficients of the covariates, here treated as nuisance parameters. Suppose that the total variance is decomposed to a genetic variance and remaining residual variance. Let 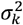 be the genetic variance component, and 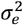 be the residual variance component, so that

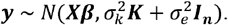

The narrow-sense heritability, defined as the proportion of total variance explained by additive genetic factors is:

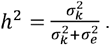

Here, we assume, without loss of generality, that 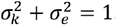. Therefore, we have 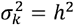, meaning that the genetic variance is equal to the heritability. Under this assumption, the variance of the phenotype can be written as

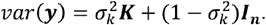

Given two *n* × 1 vectors ***y***_**1**_,***y***_**2**_, their covariance can be modelled as

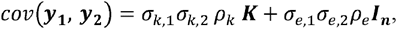

where 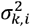 is the genetic variance for phenotype 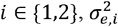 is the residual variance for phenotype *i, ρ*_*k*_ is the genetic correlation between the two phenotypes, and *ρ*_*e*_ is the residual correlation between the two phenotypes (15). Alternatively:

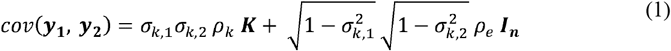

If we further plug in 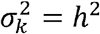, then, for a single and for two phenotypes, we can write the variance model as:

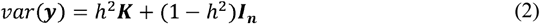

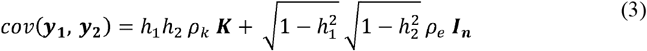

which is the form that we will use to simulate outcomes in the following parametric bootstrap section.

### Parametric Bootstrap

We use a parametric bootstrap approach to compute confidence intervals. In brief, we simulate data for each set of potential values of heritabilities 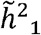,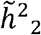 and genetic and residual correlation 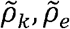 between two phenotypes based on the existing genetic relationship between individuals in the dataset of interest. Next, we compute confidence intervals by inferring ranges of potential values of *ρ*_*k*_ (integrated over potential values of 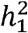,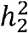, as the true values are not known) that resulted in realized (estimated) values 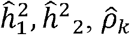. In practice, to limit computational burden, we fix 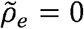 (and we assess the use of 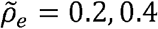 also as fixed values in simulations).

**Step 1: Random sampling of genetically correlated outcomes**

For every given combination of the potential heritability of phenotype 1 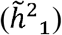, potential heritability of phenotype 2 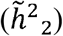, and potential genetic correlation 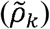, we draw *N* (e.g., 10,000) pairs of phenotype vectors (***y***_**1**_, ***y***_**2**_) from the multivariate normal distribution

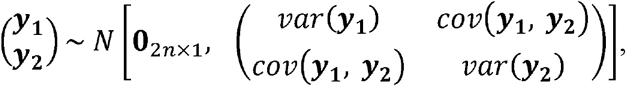

where

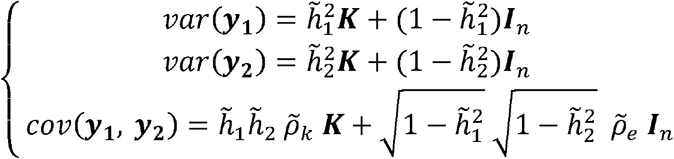

where 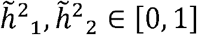 and 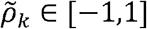. We note here that *ρ*_*e*_ may take potential value in the interval [−1,1], but we choose just one value as mentioned earlier. We used 10 settings for 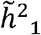, 10 settings for 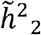, and 20 settings for 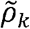 as follows:

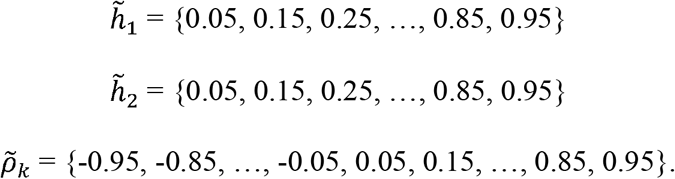

Under this setup, there are 2,000 distinct combinations of triplets 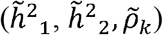 in total. We note that while developing this procedure we compared using finer grids of values, with sequences with differences of 0.01 between each two consecutive values, but the results remained essentially the same while the computational burden was substantially higher. Because the grid size determines the accuracy level of potential confidence interval coverage, we later offer a solution to increase coverage without simulating a finer grid of values.

**Step 2: Genetic Correlation and Heritability Estimation**

Next, based on each sampled pair of phenotype vectors (***y***_**1**_,***y***_2_) we estimate 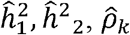. While the procedure is in principle naïve to the specific formula used, we are using the closed-form Hasemen-Elson formulas we previously derived (15,20):

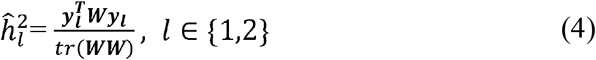

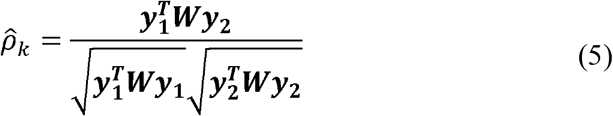

Where ***W*** is either the kinship matrix with all diagonal values set to zero, or, a weighted sum of the kinship matrix ***K*** and the matrix modelling the random error (here, an identity matrix) with weights related to the relationship between the kinship matrix and the identity matrix. See (15) for more details, including the potential use of multiple matrices modelling correlations between individuals. In practice, it is appropriate to use the kinship matrix with diagonal value set to zero when only the kinship matrix is used to model relationship between individuals. Using these formulas rather than likelihood-based procedures is computationally quicker as no iterations are required.

**Step 3: PMF estimation for** 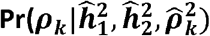)

We now derive the expression for the conditional probability of *ρ*_*k*_ given the estimated parameters. Because the support of 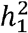,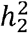,*ρ*_*k*_ are continuous where *h*_1_,*h*_2_ ∈ [0,1] and *ρ*_*k*_ ∈ [-1,1], while the results from simulations are discrete values, we divide these ranges into bins, e.g., of size 0.1, i.e., forcing them into a discrete distribution:

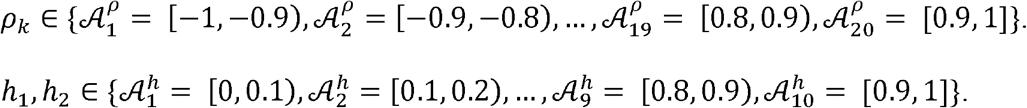

When estimating CIs for genetic correlations, we are given the estimates 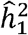,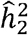,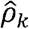 and we want to identify a region 𝒜 such that 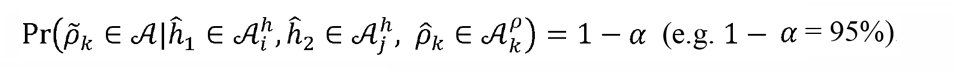. Therefore, we want to estimate the probabilities 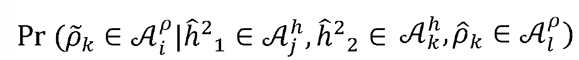 for *i* = 1, …,20 in order to create an empirical probability mass function (PMF) and use it to generate confidence intervals, which can be derived using Bayes theorem. The derivation below uses the probabilities 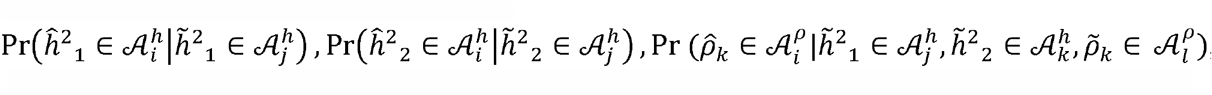, which are the probabilities of 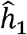,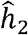,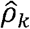 being in given regions conditional on the fixed values of 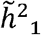,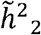,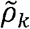 (note that the probabilities of the estimated heritabilities do not depend heritabilities of other traits on of the genetic correlation between them). Moving forward, we drop the notations showing that values refer to bins (regions) for brevity, with the understanding that all probabilities refer to parameters being in bins. Therefore, we will denote 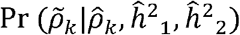 instead of 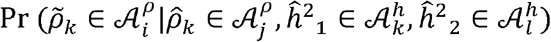, etc.

We first note that 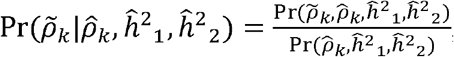, and therefore, we need to estimate 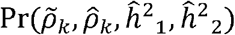 and 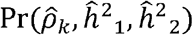.

### Estimating 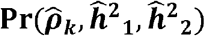

We estimate 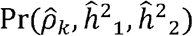 based on the following:

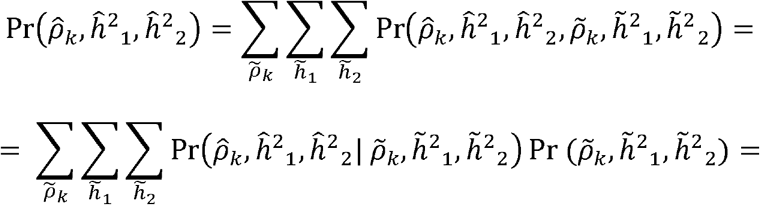

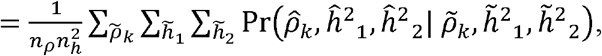

where, for bins of length 0.1, *n*_*p*_ = 20, *n*_*h*_ = 10.

### Estimating 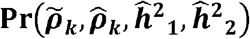

We estimate 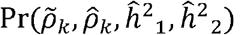 based on the following:

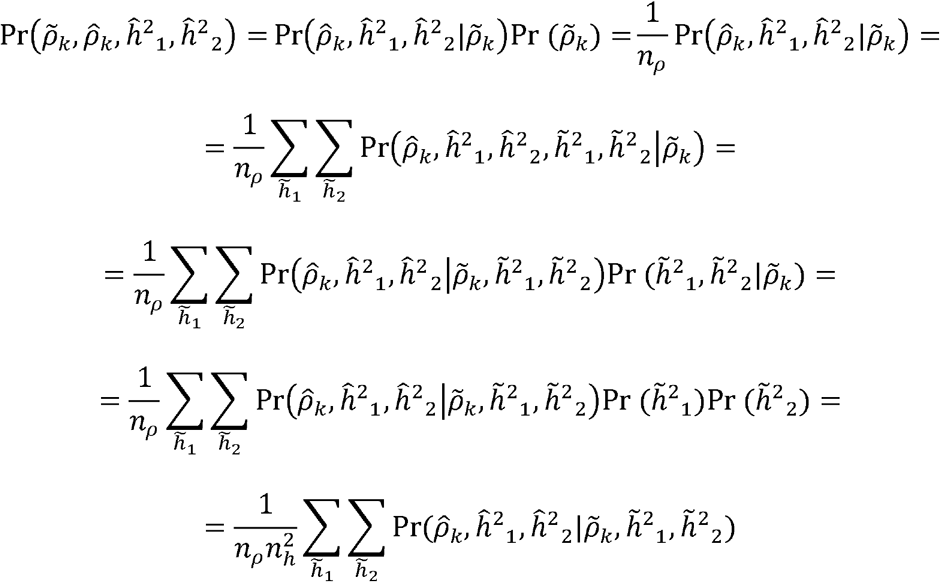

Putting these together:

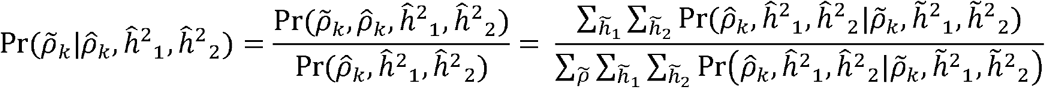

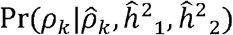 (computed for each pre-defined region) is then the empirical probability mass function of *ρ*_*k*_ obtained by parametric bootstrap.

### Computing confidence intervals from the PMF

After obtaining the empirical PMF from parametric bootstrap, we can now derive the CIs for any given genetic correlation estimate 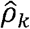 with a coverage probability of 1 − *α* (e.g., 95%). Because the parameters are bounded, constructed confidence intervals may be asymmetric in both the distance between the estimated 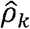 to the low and high values of the confidence interval, and in the cumulative probability between provided by the two “sides” (around 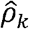) of the confidence interval. We address this by considering the following three cases depending on the cumulative probability

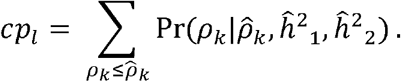

Here *cp*_*l*_ denotes cumulative probability of potential *ρ*_*k*_ values lower or equal to the estimated 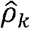. Denote the low and the high values of the confidence interval for 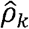 by 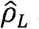 and 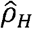. Then a 1-*α* confidence interval news to include all potential values 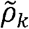 such that:

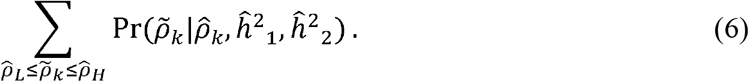

**Case 1:** If 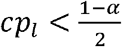

Here, 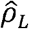 corresponds to the first potential value 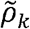 (i.e. a point in the first considered bin, where bins are considered by order 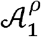,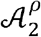 …) where 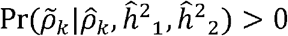.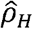 corresponds to the smallest potential value 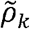 satisfying equation (6).

**Case 2:** If 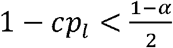

In this case we first identify 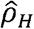 as the highest 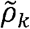 (i.e. a point in first considered bin, where bins are considered by order 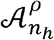,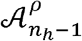 …) with 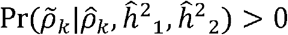.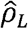 corresponds to the highest potential value 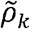 satisfying equation (6).

**Case 3:** Both 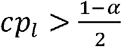 and 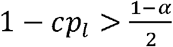

Here we require 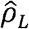 to be the largest value and 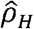 to be the lowest such that

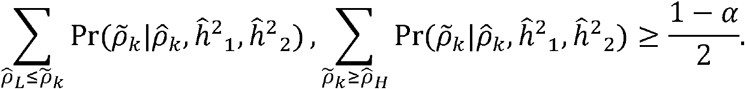

When the upper bound or lower bound of CIs obtained from the above procedure lies somewhere inside the bins defined by the grid of considered values, which is often the case, we use linear interpolation to get a position for upper and lower bound as point within the bins.

### Empirical Beta Approximation to the PMF for CI estimation

Because the PMF is discrete, it limits the potential coverage of constructed CIs and the potential computation of accurate p-values. Thus, we study a continuous beta approximation to the empirical PMF from parametric bootstrap. Since the range of genetic correlation is [-1, 1], and the range of beta distribution is [0, 1], we first map the [-1, 1] range of genetic correlations to [0,1] range of beta distribution through a location-scale transformation. After finding the 100*(1 − *α*)% CIs of 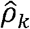 on the beta scale using a similar approach to that reported based on the discreate PMF, we apply the inverse location-scale transformation from [0, 1] to [-1, 1] to retrieve the CIs of genetic correlations.

### The Jackson Heart Study

The Jackson Heart Study (JHS) is a longitudinal study following 5,306 individuals of African American background from Jackson Mississippi (25,26). The study population included 2,050 related and unrelated JHS participants who had whole genome sequencing (WGS) through the Trans-Omics for Precision Medicine (TOPMed) program, proteomics data, and available body mass index (BMI). The TOPMed Data Coordinating Center used TOPMed WGS data from the TOPMed freeze 8 release and computed kinship matrix, tabulating the genetic relationship between TOPMed participants. We subsetted this matrix into JHS participants. Concentration levels of 1,317 proteins were measured using the SomaScan platform (27). The JHS study was approved by Jackson State University, Tougaloo College, and the University of Mississippi Medical Center Institutional Review Boards, and all participants provided written informed consent.

We excluded 5 proteins with more than 80% missing values. The remaining dataset had no missing protein measurements. Protein measurements were adjusted for batch effect by rank-normalizing each protein separately in each batch and then aggregating the data across batches. Next, the protein measurements were regressed over (1) only intercept, and (2) over age, sex, and BMI. The residuals from each of these regressions were extracted and were used for estimating heritabilities and genetic correlations between all protein pairs using Haseman-Elston estimators provided in equations (4) and (5), in addition to heritabilities and genetic correlations estimated using the rank-normalized protein levels (without regressing them on covariates). Also, we compared the estimates of genetic correlations to estimated Pearson correlations calculated using *stats* R package (version 3.6.2).

### Simulation Studies

We used the kinship matrix from the JHS data to perform simulations. To study methods’ performance in larger sample sizes, we also created simulated datasets mimicking the JHS in which we used block matrices, with blocks being the original JHS kinship matrix using n = 2,050 individuals. We used 2 and 3 times the original sample size to form block diagonal kinship matrices with n = 4,100 and n = 6,150. We referred to simulations using the kinship matrix, and the 2- and 3-times block matrices as Setting A, B, and C. Thus, we used these kinship matrices to (1) perform simulations for the parametric bootstrap procedure, where in the primary we fix 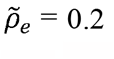 as a conservative potential high value of *ρ*_*e*_. We also performed simulations comparing the choice of 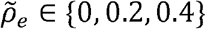. Next, (2) we generate new simulated data that used the results of the parametric bootstrap simulations (1) to construct CIs. We performed 10,000 simulations for each combination of potential 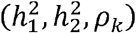, with 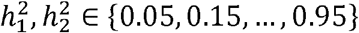 and 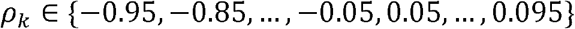. We constructed CIs for the estimated 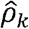 in each simulation.

#### Comparison: four approaches of constructing CIs

We estimated the coverage and the width of the CIs constructed using a few approaches: (a) percentiles of the empirical PMF constructed using the parametric bootstrap approach; (b) beta approximation to the empirical PMF; and two existing methods: (c) Fisher’s transformation; and (d) normal approximation to the distribution of the estimated genetic correlation implemented in the GCTA package (29).

The Fisher’s transformation method assumes that genetic correlations follow the same distributions as Pearson correlations (30). More specifically, they are normally distributed after Fisher’s transformation. For genetic correlation *ρ*_*k*_,

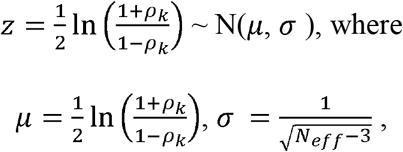

With *N*_*eff*_ being the “effective sample size”, previous proposed to be *trace*(***K***^−^***K***^−^) with ***K***^−^ being the kinship matrix with diagonal values set to zero (15). We can then construct the CIs of z by the standard approach assuming asymptotic normal distributions. For example, the 95% CI of z would be [*μ* − 1.96 *σ, μ* +1.96*σ*]. After finding the 100*(1 − *α*)% CIs of 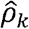 on the Fisher’s transformed (*z*) scale, we apply the inverse Fisher’s transform to retrieve the CIs of the genetic correlation *ρ*_*k*_.

To compute CIs based on existing approach that rely on a normal approximation, we estimate both the genetic correlation and its standard error using the bivariate REML procedure implemented in the GCTA package. We apply the 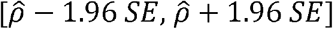 formula to construct 95% CIs. Due to the high computational resources required by GCTA, we focus only on the four scenarios when true *ρ*_*k*_ equals {0.05, 0.15, 0.45, 0.95} with the original-size kinship matrix.

#### Performance evaluation metrics

We used coverage probabilities and CI widths as the metrics to evaluate and compare the performance of the four approaches for CI construction. In primary results, for a given true value of genetic correlation *ρ*_*k*_ we calculated both the coverage probability and the average width of 95% CIs using the constructed CIs for the estimated *ρ*_*k*_ over all the 100 true heritability scenarios (10 for 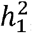, and 10 for 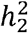). Ideally, the coverage probabilities would be at or above 95% across different *ρ*_*k*_, and also having small CI widths. The coverage probability was estimated as the proportion of simulations in which the true *ρ*_*k*_ was contained in its CI.

#### P-value estimation

We evaluated the use of the CI inversion method to obtain p-values for hypothesis testing. Here,our null hypothesis H_0_ is *ρ*_*k*_ = 0, and our alternative hypothesis H_1_ is *ρ*_*k*_ ≠ 0. Given any realization 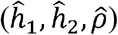, we can estimate the CIs for *ρ*_*k*_ using the parametric bootstrap procedure, focusing on the continuous beta approximation to the empirical PMF because smaller accurate p-values can be obtained if the underlying distribution is continuous. To determine the p-values of genetic correlation estimates, we use the CI inversion methods. Suppose that we construct a 100× (1 − *α*)% CI. Then, we can determine that the p-value is smaller than *α* if the constructed CI does not cover 0. For computational efficiency, we implemented a method that constructs CIs using a binary search approach to the *α* value, stopping when a pre-defined sensitivity level is reached.

#### Type 1 error

For each combination of potential heritability values 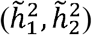, we simulated 10,000 pairs of phenotype vectors (***y***_**1**_***y***_**2**_) under the null, i.e., *ρ* = 0, estimated their genetic correlations, and calculated their p-values as described above based on the beta approximation to the PMF. After obtaining the p-values for all the 10,000 simulated data, we estimated the type 1 error rate, also called the size of the test, by checking the percentage of these simulation rejecting the null given an *α* value.

## Results

### Simulation studies

In primary results, for a given true value of genetic correlation *ρ*_*k*_ we calculated both the coverage probability and the average width of 95% CIs by averaging the corresponding estimates over all the 100 true heritability scenarios (10 for 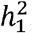, and 10 for 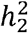). Supplementary Table 1 provides estimated coverage by all combinations of (true) ρk,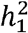,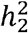. Figure 1 provides the estimated coverage probabilities for the compared methods in simulations, and Figure 2 provides the averaged CI widths. The PMF approach provides appropriate coverage across the three settings defined by the kinship matrices. The beta approximation to the PMF results in under-coverage across the simulated *ρ*_*k*_ values setting A, but improved substantially in settings B and C when the simulated sample size increased. Still the average width of the CIs was lower when using the empirical PMF. In setting A, GCTA had an appropriate coverage only when *ρ*_*k*_ was set to 0.05. The Fisher’s transformation tended to result in under-coverage.

**Figure 1.**
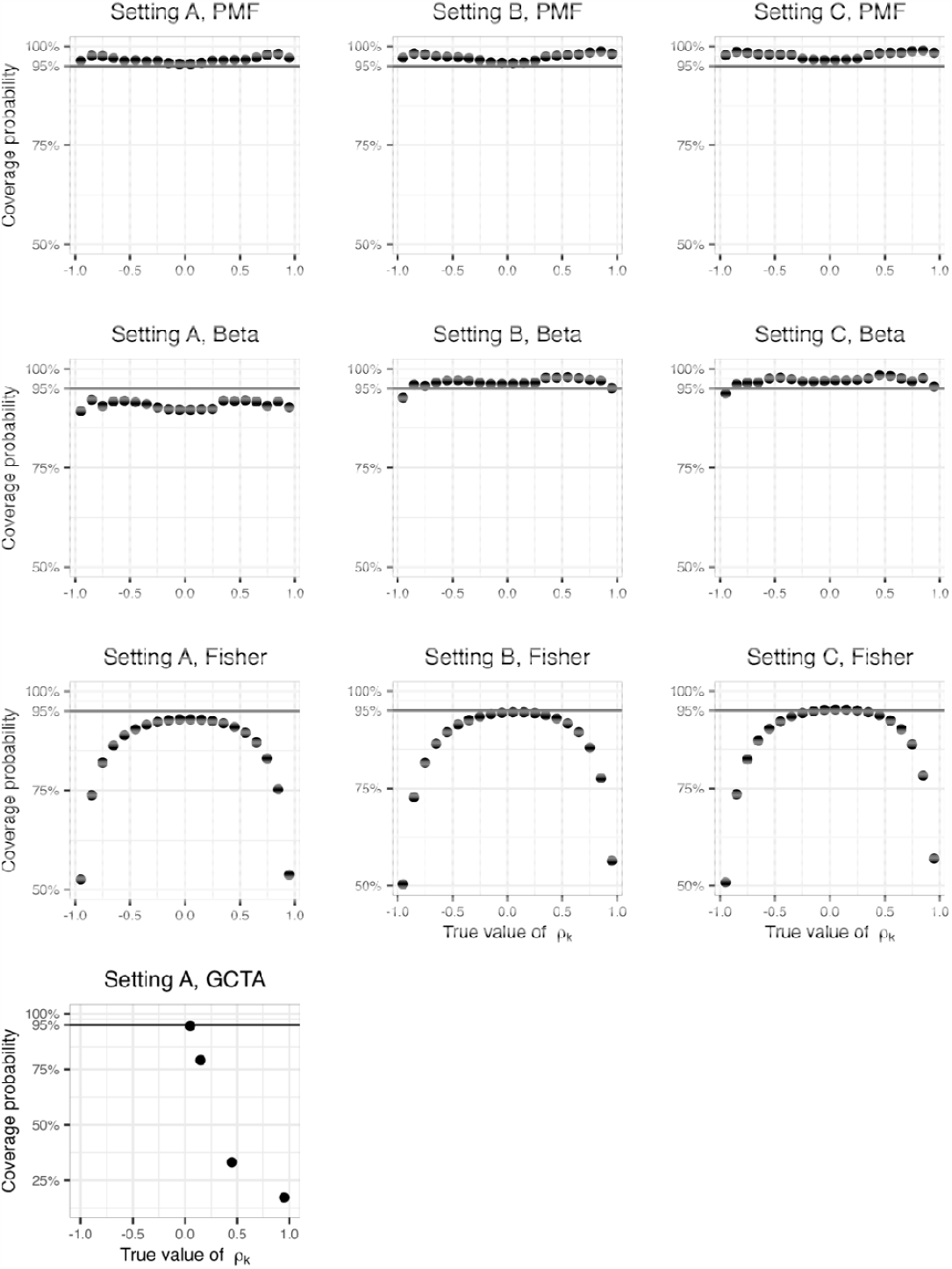
Estimated coverage of 95% confidence intervals of genetic correlations in the primary simulations. The columns represent different kinship matrix sizes: Setting A denotes the use of original-size kinship matrix (n=2,050), Setting B denotes the use of the double-size kinship matrix (n=4,100), and Setting C denotes the use of the triple-size kinship matrix (n=6,150). The rows represent the four approaches for constructing CIs, including parametric bootstrap PMF, beta approximation for parametric bootstrap PMF, Fisher’s transformation, and GCTA package use of normal distribution approximation. Only parts of the analyses were carried out on the GCTA package due to the high computational resources required.

**Figure 2.**
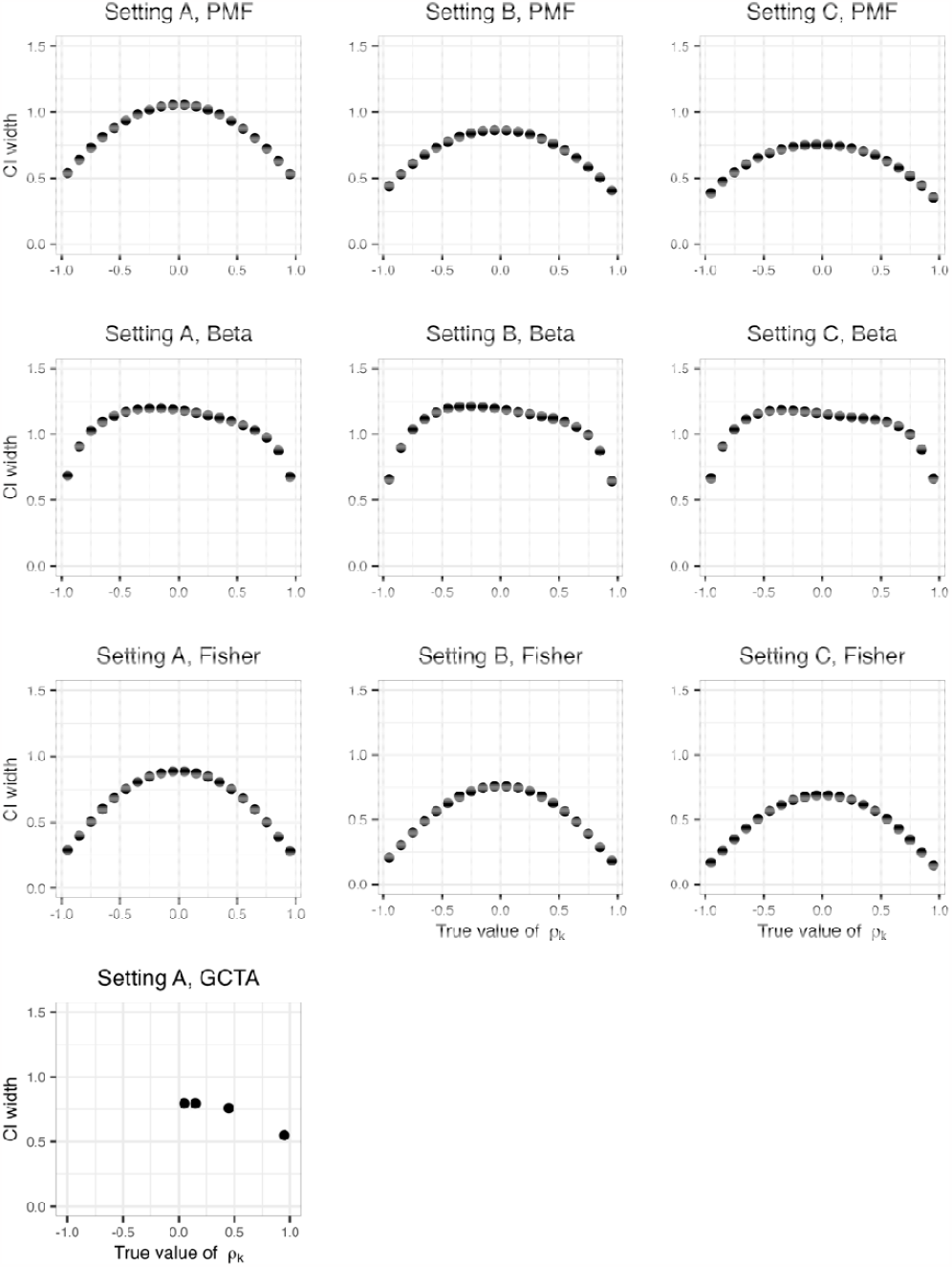
Average width of the confidence intervals of the genetic correlations in the primary simulations. The columns represent different kinship matrix sizes: Setting A denotes the use of original-size kinship matrix (n=2,050), Setting B denotes the use of the double-size kinship matrix (n=4,100), and Setting C denotes the use of the triple-size kinship matrix (n=6,150). The rows represent the four approaches for constructing CIs, including parametric bootstrap PMF, beta approximation for parametric bootstrap PMF, Fisher’s transformation, and GCTA package. Only parts of the analyses were carried out on the GCTA package due to the high computational resources required.

Figure 3 compares coverage and CI widths when using the empirical PMF approach to compute CIs and settings *ρ*_*e*_ = 0,0.2, or 0.4 in the simulations generating data. It demonstrates that there is essentially no difference in the results.

**Figure 3.**
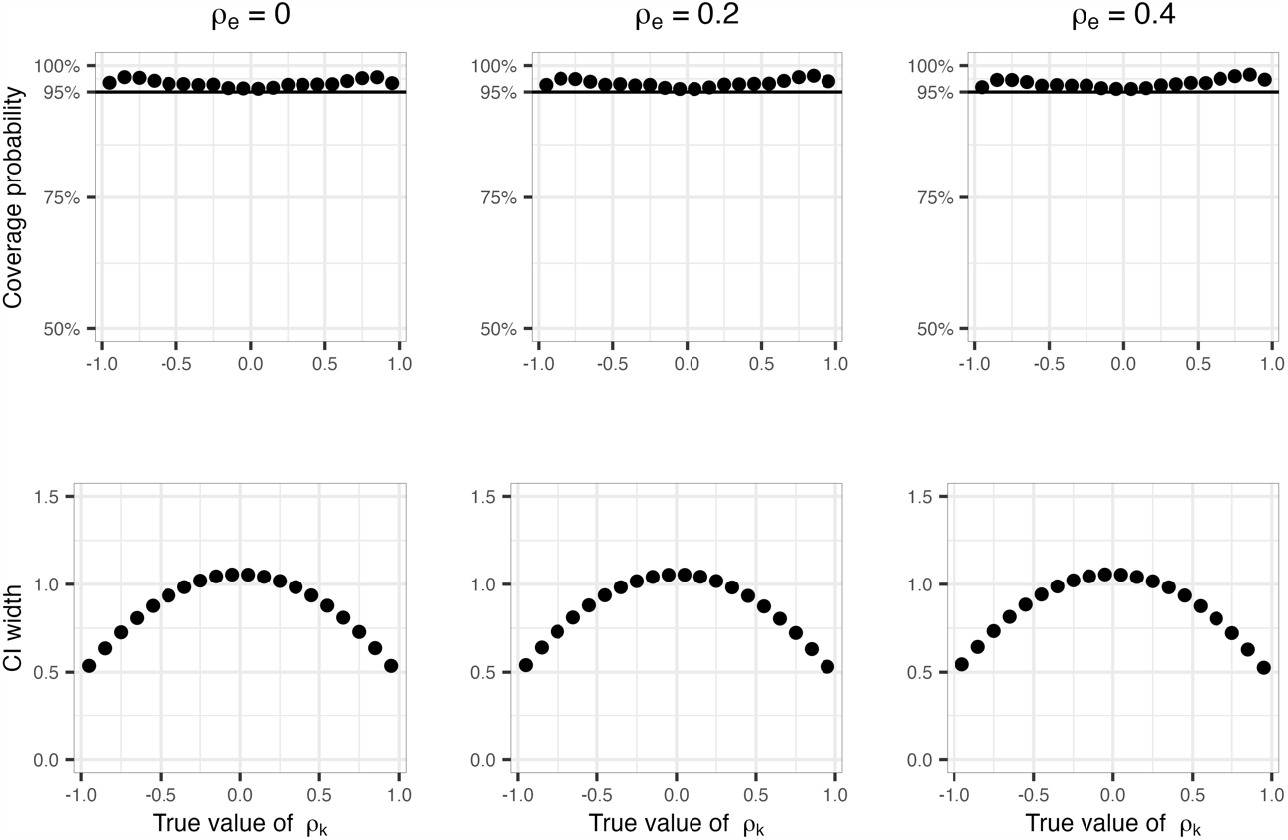
Setting environmental correlation between error terms has no effect on genetic correlation estimates. Comparison of coverage and CI widths when using the empirical PMF approach to compute CIs and settings or in the simulations generating data.

We also used the simulations to estimate type 1 error when using the confidence interval inversion methods with the beta approximation to the PMF to compute association p-values. The results are visualized in Figure 4. Here, we also estimated type 1 error by combinations of specific 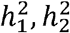 values. With *α* = 0.05, the type 1 error was controlled across heritability combinations in settings B and C, but not in setting A. While it is unsurprising that the type 1 error is not controlled when heritability values of either one of the two traits are very small, the test was also somewhat inflated in setting A when the two heritabilities were fairly high. Over all, the beta approximation method to the PMF is promising for computing high coverage CIs and p-values when the sample size is sufficiently large.

**Figure 4.**
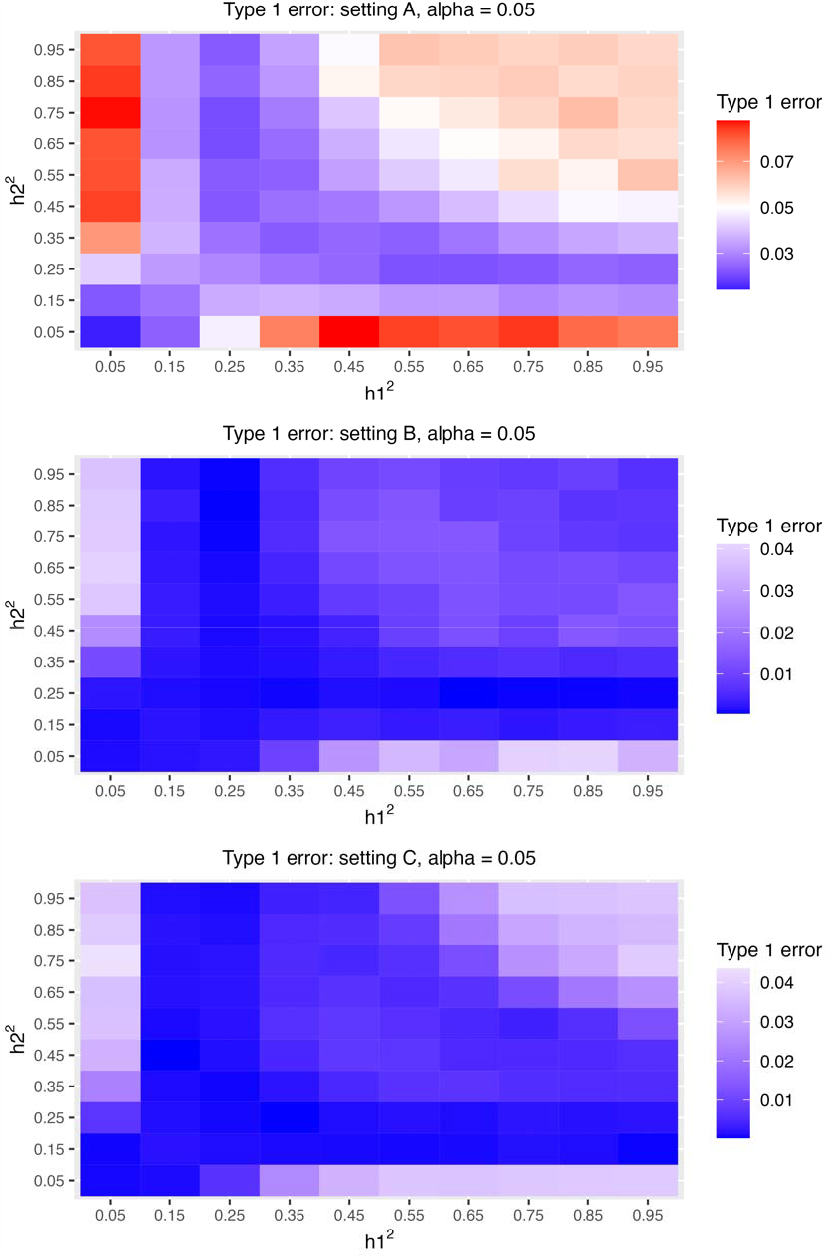
Type 1 error estimates when using the confidence interval inversion method and the Beta approximation to perform association testing. Visualization of type 1 error () when using the CI inversion approach coupled with the Beta approximation to the PMF to generate CIs. The results are provided for each simulation settings and by values of.

### Application to genetically-determined protein-protein networks in JHS

We estimated heritabilities and genetic correlations for every pair of proteins among the 1,317 proteins available in JHS, in an analysis adjusted to age, sex, and BMI (in which protein measures were first regressed over these covariates prior to estimation of genetic correlations based on the resulting residuals), and in an unadjusted analysis. Characteristics of the JHS dataset are provided in Table 1. Of the study participants, 61% were women. Individuals were 55 (male)-56 (female) on average, and were mostly overweight. Some individuals were close. For example, there were 341 pairs of individuals with estimated coefficient of relationship ≥ 0.48, and 1,113 pairs of individuals with coefficient of relatives relationship ≥ 0.12 (considering the total number of unique pairs of individuals, this corresponds to 0.05% of all pairs of participants).

**Table 1:**
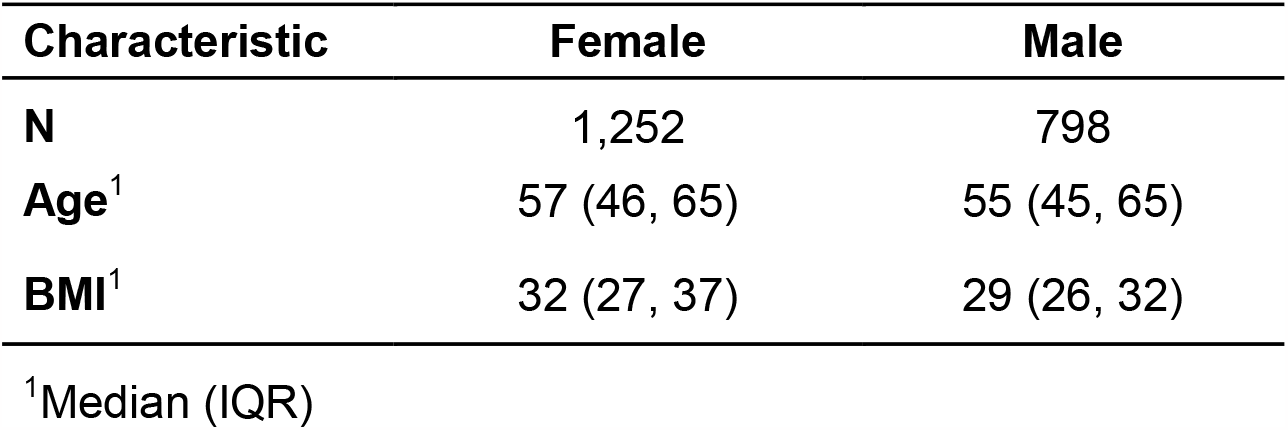
JHS dataset characteristics stratified by sex.

Supplementary Tables 2 and 3 provide the estimated heritabilities of all proteins in the dataset from analysis unadjusted and adjusted to covariates (age, sex and BMI) respectively. Based on the simulations using this specific dataset, we removed from consideration proteins with estimated heritabilities 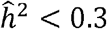, as genetic correlations and p-values using the beta, approximation method are less reliable compared to higher values of (real, not estimated) heritabilities. We also excluded proteins with estimated 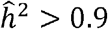 because such high may suggest a problem with the measurement and/or genetic characterization of a protein (e.g., technical issue with the platform, genetic variants segregated to a few families, etc.). After the above filtering, there were 403 and 431 proteins, or 81,406 and 93,096 protein-protein pairs, available for genetic correlation analysis in the covariate-adjusted and unadjusted analyses, respectively. For each set of the proteins (adjusted and unadjusted), it took around 2.5 hours to estimate the genetic correlations and around 12 minutes to construct the CIs, based on the previously-constructed parametric bootstrap reference results, for all the protein-protein pairs on a MacBook Pro laptop with an M1 chip. Full results from genetic correlation estimates for these sets of proteins are provided in Supplementary Tables 4 and 5. Figure 5 visualizes the comparison between estimated phenotypic (Pearson) and genetic correlations across these phenotype pairs. The figure suggests that, for this set of highly-heritable proteins, genetic correlations tend to be higher than Pearson correlations (to see this, one needs to focus in Figure 5 on the bright hexbins because they represent many more protein pairs compared to dark hexbins).

**Figure 5.**
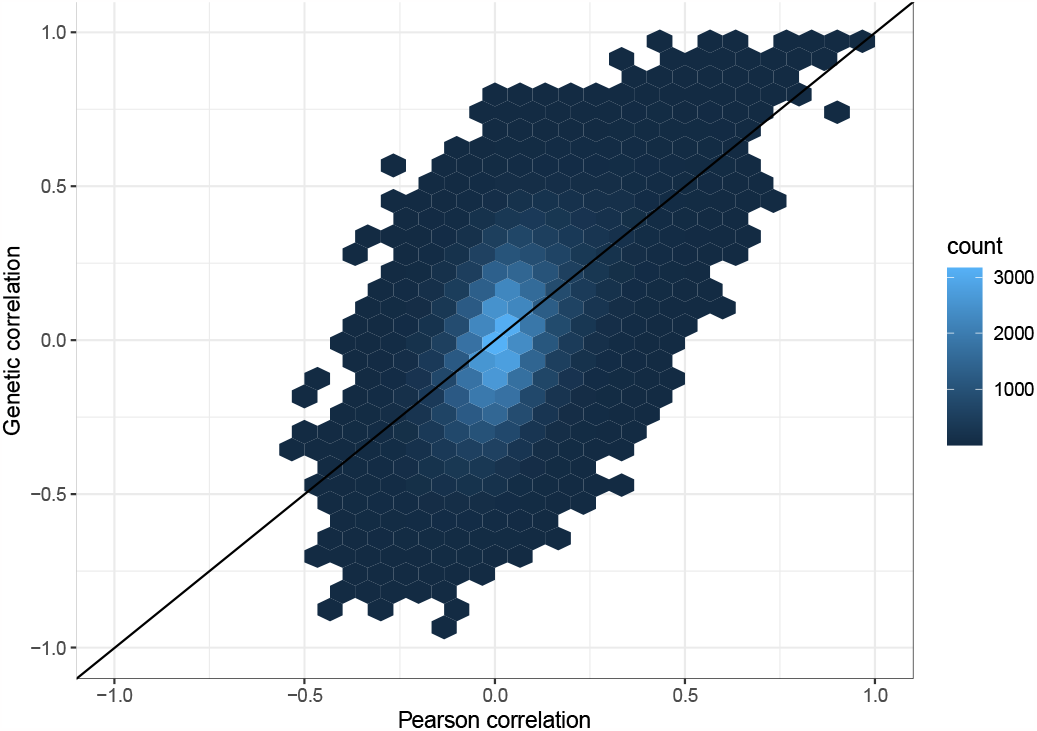
Estimated Pearson versus genetic correlations between heritable proteins. The figure compares the sample Pearson correlation to the estimated genetic correlation 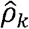for all protein pairs for which the estimated heritability 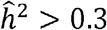 for each of the proteins. The color of each hexbin represents the number (count) of protein pairs with x- and y-axis values falling under the hexbin.

#### Protein-Protein Network

We visualize the results in a protein-protein network. Due to the large number of protein pairs, we focused the network resulting from protein-protein genetic correlations passing a p-value threshold. We computed p-values for the genetic correlations between the limited set of heritable and “valid” proteins (with heritability estimates that are not egregiously high) using the beta approximation to the PMF, and applied a False Discovery Rate (FDR) correction using the Benjamini-Hochberg procedure (31). The considered pairs of proteins are those with FDR-adjusted genetic correlation p-value<0.01. This corresponds to 253 and 294 pairs of genetically-correlated proteins in adjusted and unadjusted analysis, respectively. Figure 6 visualizes these results. The size of each node represents its degree, with larger ones being “hub nodes/proteins”, (genetically) associated with a large number of proteins. See Supplementary Tables 6 and 7 for estimated genetic correlations between pairs of proteins selected based on the criteria described above. Supplementary Table 8 contains a list of the top 10 hub nodes/proteins and their connections, i.e., the list of proteins connected to each of these hub nodes, both in covariate adjusted and unadjusted analyses. Visually, the network appears to be less connected (and we also know that the number of connections decreased) in analysis that adjusted for age, sex, and BMI. It is likely that genetic correlations decreased because BMI has strong effects on proteins, and the genetic effects on BMI are also strong, so when BMI was adjusted for, genetic effects inducing correlations between proteins were reduced.

**Figure 6.**
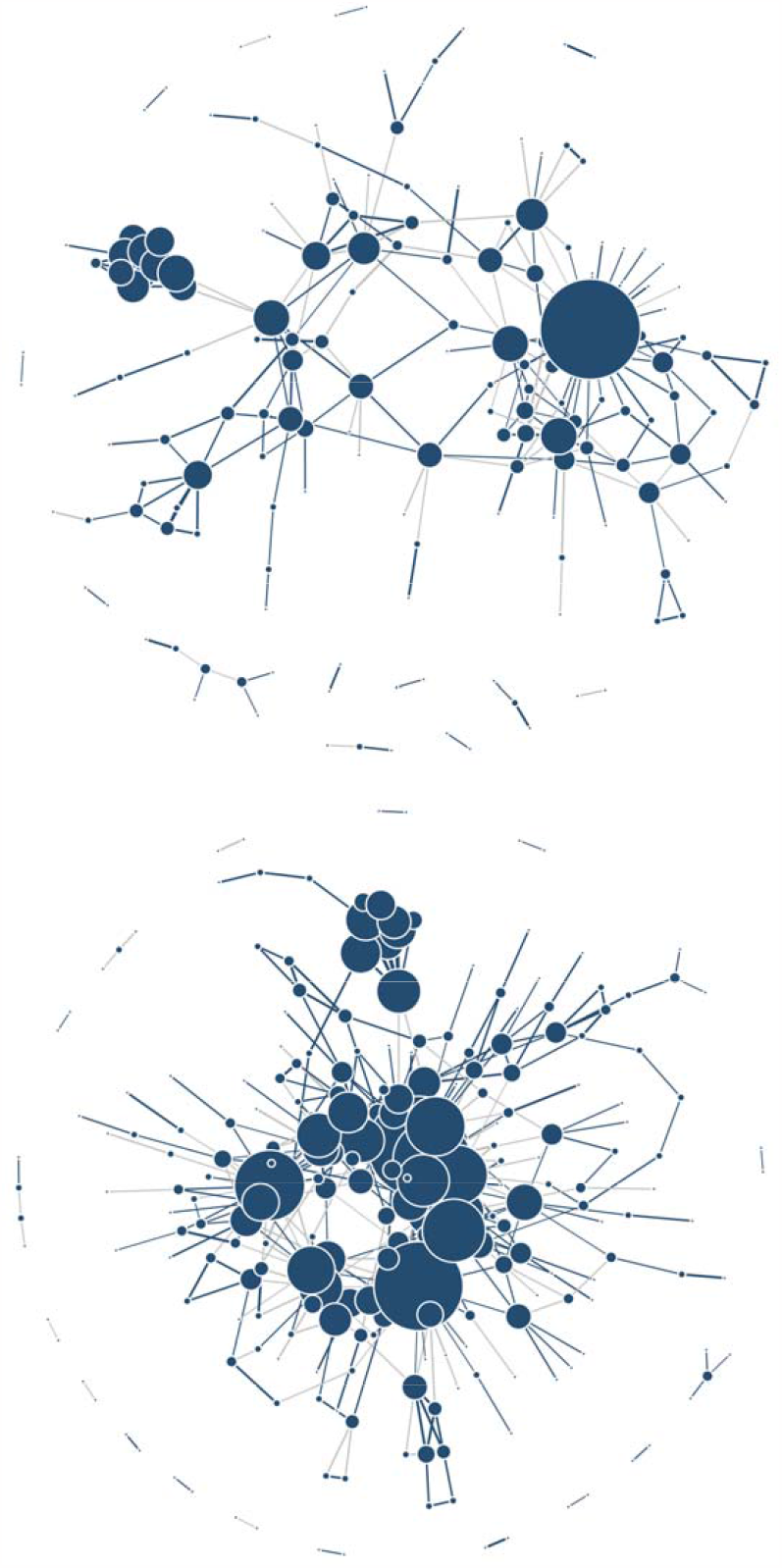
Network constructed from top pairs of genetically-correlated proteins. Panel (a) visualizes the protein-protein genetic correlation network using the age, sex, BMI-adjusted proteins; panel (b) visualizes the corresponding network based on unadjusted analysis. The blue edges represent positive genetic correlations, and the grey edges represent negative genetic correlations. Larger nodes are hub proteins where multiple proteins have strong genetic correlations with each other both in covariate adjusted and unadjusted analyses. Some of the hub proteins include *APO_D, PREKALLIKREIN, NOTCH_3, HPV_E7_TYPE18, CARBONIC_ANHYDRASE_IV, CDK5_P35, DKK_4, PAK3, TRKC, MIS, C5A, OMD, JAG1, HEPARIN_COFACTOR_II, BFGF_R, and MMP_2, GDF_11_8*.

## Discussion

We developed a parametric bootstrap procedure to estimate confidence intervals for the genetic correlation estimator, studied it in simulations, and applied it to learn a protein-protein network using a set of heritable proteins measured in the Jackson Heart Study. Our bootstrap procedure was inspired by a similar approach developed for heritability confidence intervals (19).

Compared to the previous publication focusing on heritability, our approach is complicated by the need to simulate pairs of traits, including their heritabilities and genetic correlation between them, i.e., a grid of three parameters rather than one. Indeed, confidence intervals for genetic correlation depend on trait heritability, and are wider when at least one of the traits has low heritability. Thus, the computation burden of our procedure is higher. Especially, it is important to recognize that this procedure, like that of Schweiger et al., is dataset dependent, because it uses the kinship matrix of the specific dataset. However, our procedure is realistic and useful when many genetic correlations are estimated for the same dataset, as in this work. In this case the parametric bootstrap simulation step is performed once but is applied many times. A limitation for the high dimensional number of parameters (many genetic correlation parameters) is the limited level of coverage due to the discreteness of the bootstrap procedure: we cannot use the estimated conditional PMF of *ρ*_*k*_ (conditional on the estimated genetic correlation and heritabilities) as it is to obtain confidence intervals at the 1-*α* level when *α* is very small (e.g., 10^−7^). To address this, we proposed the beta approximation, after local-scale transformation, to the PMF. The beta distribution has two parameters that can be fit to many distribution functions that are on a bounded interval. Based on our simulations, CIs based on the beta approximation tend to be wider than those using the PMF directly, and they can still undercover the desired distribution in low sample sizes. However, for larger sample sizes their performance improves. Overall, we think that for larger sample sizes, e.g., 6,000 individuals, the beta approximation to the PMF will be very useful in providing reliable confidence intervals and, using the inversion method, p-values. It is important to point out that while we performed simulations with a “triple size” JHS kinship matrix, i.e., of n=6,150 individuals, the effective sample size corresponding to it is much lower than that of real potential datasets with 6,150 individuals. That is because we simulated a block diagonal matrix. Realistic kinship matrices will have non-zero off diagonal values throughout (unless forced to be zero for computational efficiency purposes (32)).

Existing methods that compute confidence intervals for genetic correlations typically utilize an asymptotic normal distribution argument, at either the untransformed or Fisher’s transformation level. This is appropriate depending on the combination of four factors: sample size, underlying (true) heritabilities of each of the pair of traits, and the underlying genetic correlation. For any given pair of traits and a dataset, any one of these factors may be suboptimal, potentially leading to poor performance of confidence intervals that rely on asymptotic normality. The bootstrap procedure addresses this shortcoming. However, this procedure too does not produce perfect confidence intervals: for low values of heritability of either one of the two traits, the coverage may still be lower than desired in low sample sizes. Note that in reality we do not know the true heritability, we only have estimated heritability. Therefore, we cannot tell whether a CI may not be reliable according to the values of the estimated heritabilities. That is why our main results are provided at an aggregate level, across simulated values of potential heritabilities.

We demonstrated the use of genetic correlations to infer genetically-determined protein-protein networks. However, we acknowledge that our analysis is limited by the relatively low sample size, which led to posing a stringent filter requiring at least 0.3 protein heritability for inclusion in the downstream analysis. While we chose to include only edges with estimated FDR-adjusted p-value<0.01 (with p-values estimated using the beta approximation), other statistical network approaches may generate sparsity using penalized multivariant regression techniques (33,34). It would be interesting to extend such approaches to genetic (rather than phenotypic) correlation-based networks. In future work we will apply the existing framework on larger datasets and develop approximation methods to further speed up the simulations required for the parametric bootstrap and the estimation of heritabilities and genetic correlations, for example, following (35).

## Data Availability

Individual-level JHS data can be obtained by application to dbGaP (accession phs000286) or by data use agreement with JHS Data Coordinating Center (DCC), see study website at https://www.jacksonheartstudy.org/. Summary statistics from analyses reported in this manuscript are provided as supplementary materials.

## Acknowledgements

This work was supported by the National Institute of Diabetes and Digestive and Kidney Diseases R01DK081572. The Jackson Heart Study (JHS) is supported and conducted in collaboration with Jackson State University (HHSN268201800013I), Tougaloo College (HHSN268201800014I), the Mississippi State Department of Health (HHSN268201800015I) and the University of Mississippi Medical Center (HHSN268201800010I, HHSN268201800011I and HHSN268201800012I) contracts from the National Heart, Lung, and Blood Institute (NHLBI) and the National Institute on Minority Health and Health Disparities (NIMHD). The authors also wish to thank the staffs and participants of the JHS. The views expressed in this manuscript are those of the authors and do not necessarily represent the views of the National Heart, Lung, and Blood Institute; the National Institutes of Health; or the U.S. Department of Health and Human Services.

